# Large Individual Differences in Functional Connectivity in the Context of Major Depression and Antidepressant Pharmacotherapy

**DOI:** 10.1101/2023.10.17.23297087

**Authors:** Gwen van der Wijk, Mojdeh Zamyadi, Signe Bray, Stefanie Hassel, Stephen R. Arnott, Benicio N. Frey, Sidney H. Kennedy, Andrew D. Davis, Geoffrey B. Hall, Raymond W. Lam, Roumen Milev, Daniel J. Müller, Sagar Parikh, Claudio Soares, Glenda M. Macqueen, Stephen C. Strother, Andrea B. Protzner

**Affiliations:** University of Calgary, Department of Psychology, Calgary, Canada; Rotman Research Institute, Baycrest Health Sciences, Toronto, Canada; Child and Adolescent Imaging Research Program, University of Calgary, Calgary, AB, Canada; Alberta Children’s Hospital Research Institute, University of Calgary, Calgary, AB, Canada; Hotchkiss Brain Institute, University of Calgary, Calgary, AB, Canada; Department of Radiology, University of Calgary, Calgary, AB, Canada; University of Calgary, Cumming School of Medicine, Department of Psychiatry, Calgary, Canada; Mathison Centre for Mental Health Research and Education, University of Calgary, Calgary, Canada; Department of Psychiatry and Behavioural Neurosciences, McMaster University, Hamilton, Canada; Mood Disorders Program and Women’s Health Concerns Clinic, St. Joseph’s Healthcare, Hamilton, Canada; Department of Psychiatry, University of Toronto, Toronto, Canada; Institute of Medical Sciences, University of Toronto, Toronto, Canada; Centre for Mental Health, University Health Network, Toronto, Canada; Centre for Depression and Suicide Studies, Unity Health Toronto, Toronto, Canada; Krembil Research Institute, Toronto Western Hospital, Toronto, Canada; Department of Psychology, Neuroscience & Behaviour, McMaster University, Hamilton, Canada; Imaging Research Centre, St. Joseph’s Healthcare Hamilton, Hamilton, Canada; University of British Columbia, Department of Psychiatry, Vancouver, Canada; Queen’s University, Departments of Psychiatry and Psychology, and Providence Care Hospital, Kingston, Canada; Campbell Family Mental Health Research Institute, Centre for Addiction and Mental Health, Toronto, Canada; Department of Pharmacology & Toxicology, University of Toronto, Toronto, Canada; Department of Psychiatry, University of Michigan, Ann Arbor, Michigan, USA; Department of Psychiatry, Queen’s University, Providence Care, Kingston, Ontario, Canada; University of Toronto, Department of Medical Biophysics, Toronto, Canada

## Abstract

Clinical studies of major depression (MD) often examine differences between groups whilst ignoring within-group variation. However, inter-individual differences in brain function are increasingly recognised as important and may impact effect sizes related to group effects. Here, we examine the magnitude of individual differences in relation to group differences that are commonly investigated (e.g., related to MD diagnosis and treatment response). Functional MRI data from 107 participants (63 female, 44 male) were collected at baseline, 2 and 8 weeks during which patients received pharmacotherapy (escitalopram, N=68), and controls (N=39) received no intervention. The unique contributions of different sources of variation were examined by calculating how much variance in functional connectivity was shared across all participants and sessions, within/across groups (patients vs controls, responders vs non-responders, female vs male participants), recording sessions and individuals. Individual differences and common connectivity across groups, sessions and participants contributed most to the explained variance (>95% across analyses). Group differences related to MD diagnosis, treatment response and biological sex made significant but small contributions (0.3-1.2%). High individual variation was present in multimodal association areas, while low individual variation characterized primary sensorimotor regions. Group differences were much smaller than individual differences in the context of MD and its treatment. These results could be linked to the variable findings and difficulty translating research on MD to clinical practice. Future research should examine brain features with low and high individual variation in relation to psychiatric symptoms and treatment trajectories to explore the clinical relevance of the individual differences identified here.

**Significance statement:** Studies on major depression often investigate differences in brain function between groups (e.g., those with/without a diagnosis) with the aim of better understanding this prevalent condition. Our study shows that group differences only tell part of the story, by highlighting strong common and individually unique features of brain network organization, relative to surprisingly subtle features of diagnosis and treatment success. From the overall explained variation in brain connectivity, about 50% was shared across everyone, while another 45% was unique to individuals. Only ∼5% could be attributed to diagnosis, treatment success and biological sex differences. Our results suggest that examining individual differences, and their potential clinical relevance, alongside group differences may bring us closer to improving clinical outcomes for major depression.

**Trial registration:** ClinicalTrials.gov: NCT01655706.

## Introduction

Neuroimaging studies examining major depression (MD) have identified group differences between patients and controls, for different treatment outcomes, and in relation to biological sex/gender (Bangasser & Cuarenta, 2021; Brakowski et al., 2017; Dichter et al., 2015; Gudayol-Ferré et al., 2015; Labaka et al., 2018; Mulders et al., 2015; Ruigrok et al., 2014; Wheelock et al., 2019). Although such research has increased our understanding of brain function related to MD, less attention has been given to individual differences and commonalities across groups. A recent study showed that, in neurotypical individuals, common and stable individual features were the largest contributors to the variance in fMRI functional connectivity (FC) across sessions and tasks (Gratton et al., 2018). In addition, there is evidence to suggest that such stable individual features may relate to cognition and clinical symptoms (Finn et al., 2015; Gordon et al., 2018; Wang et al., 2020). These findings highlight the potential importance of individual differences and commonalities across groups, in addition to group differences.

Several studies have investigated FC in the context of MD and antidepressant treatment (Brakowski et al., 2017; Dichter et al., 2015; Gudayol-Ferré et al., 2015; Mulders et al., 2015). Connectivity features related to MD are most consistently identified across three major resting state networks; the default mode network (DMN), salience network (SN) and cognitive control network (CCN) (Brakowski et al., 2017; Mulders et al., 2015). Similarly, connectivity features have been linked to treatment success, most commonly within the (anterior) DMN and between frontal and limbic regions (Brakowski et al., 2017; Dichter et al., 2015; Gudayol-Ferré et al., 2015). Despite such consistencies, differences across studies are also observed. Next to variable methodologies, inter-individual heterogeneity is often cited as a potential reason for inconsistencies (Brakowski et al., 2017; Müller et al., 2017).

While patients with MD are heterogeneous in clinical symptoms (Fried & Nesse, 2015) and response to treatment (Rush et al., 2006), less is known about individual differences in brain function. Instead, individual variation in the brain has mostly been investigated in neurotypical samples. These studies have identified stable individual differences (Chen et al., 2015; Gratton et al., 2018), including greater individual variation in association networks, and less in primary sensorimotor networks (Chen et al., 2015; Kong et al., 2019; Mueller et al., 2013; Seitzman et al., 2019). Importantly, this research indicates that larger amounts of data than typically used in neuroimaging studies of MD (e.g., 25-200 minutes instead of 5-10 minutes) are needed for stable individual estimates of FC (Gordon et al., 2017; Gratton et al., 2020; Laumann et al., 2015). Using longer fMRI scans (here we use 30 minutes) to explore the magnitude and spatial distribution of individual alongside group differences in the brain may therefore be especially relevant for MD, considering the hopes that neuroimaging will eventually support clinical decision making for individual patients (Fonseka et al., 2018; Fu & Costafreda, 2013).

Other relevant factors in understanding MD and treatment outcomes are biological sex, gender identity and gender expression. Though more research is needed to understand the complex biological, social, cultural, and socioeconomic causes underlying these differences, higher rates of MD have been observed in women compared to men (Whiteford et al., 2013) and the expression of MD symptoms differs between women and men (Altemus et al., 2014). Women may also respond better to SSRI’s than men (Khan et al., 2005). In terms of brain features, sex differences have been observed in both the general population (Ruigrok et al., 2014; Wheelock et al., 2019) and MD (Bangasser & Cuarenta, 2021; Labaka et al., 2018). Despite these differences, in a recent structural MRI study of neurotypical participants, individuals rarely matched the ‘typical female’ or ‘typical male’ brain, instead showing unique individual features (Joel et al., 2015), highlighting the potential importance of individual differences in this dimension as well.

In the present study, we examined the relative contribution of sources of variance in neuroimaging data in the context of MD and its treatment. To our knowledge, no other studies have quantified individual variation in psychiatric populations. We used multi-site fMRI data from the CAN-BIND initiative, including 68 patients and 39 controls (Kennedy et al., 2019; Lam et al., 2016). Participants were scanned three times (baseline, 2 and 8 weeks; ∼30 minutes of fMRI data per scan) with patients receiving escitalopram after their baseline scan. We calculated the cross-correlation between individual- and session-specific FC matrices to assess similarities and differences within and across groups (controls vs patients, treatment responders vs non-responders, female vs male participants), sessions (baseline, 2 and 8 weeks) and individuals, as well as the MD*session and MD*sex interactions. We also assessed where in the brain these effects were most prominent.

We hypothesized that considerable variance would be explained by individual differences and similarities across all participants and sessions (common effect) (Gratton et al., 2018). We expected minimal contribution from variance over sessions in controls (no change over time), and a small but significant contribution of sessions in patients (change related to treatment). Based on research examining similarities and differences between patients and controls (Winter et al., 2021) and female and male participants (Joel et al., 2015), we expected the contribution of MD diagnosis, treatment response, sex and their interactions to be significant, but smaller than the common and individual contributions. Finally, we expected the common effect to be most prominent in sensorimotor areas, individual differences to be present in multimodal association cortices (Gratton et al., 2018; Kong et al., 2019; Mueller et al., 2013), and variance explained by MD diagnosis and treatment response to be present in the DMN, SN and CCN (Brakowski et al., 2017; Dichter et al., 2015; Mulders et al., 2015).

## Methods

The data analyzed in this study were collected as part of the Canadian Biomarker Integration Network for Depression (CAN-BIND) initiative. A detailed report of the protocol [30] and primary outcomes [29] have been published elsewhere. We preregistered our analyses on the Open Science Framework (OSF; https://osf.io/79gv8/?view_only=9e105962ce4c4ebf8cf35393471a7b69), and report the changes that we made post-registration (see Transparent_Changes_Document.docx).

### Participants

The sample included in these analyses consisted of 107 participants, including 68 patients with a primary diagnosis of major depression (MD) and 39 controls. From the larger CAN-BIND-1 dataset, we selected participants who had complete and high-quality neuroimaging data for the resting state, affective go/no-go and monetary incentive delay tasks at all three fMRI recording times, to get the longest total recording times under similar conditions per participant. Sex was measured as the self-reported sex of the individual (based on biological reproductive functions) and/or the clinician’s assignment based on a physical examination and was coded as binary (female or male; see limitations section for discussion on this). Using this definition, among patients and controls respectively, 42 and 21 participants were female, while 26 and 18 were male. Participants were recruited at six sites in Canada, were 18-60 years of age and spoke sufficient English for the completion of this study. Demographic information is reported in **Table 1**. Ethics approval was granted by local ethics committees and all participants gave written informed consent.

**Table 1.**
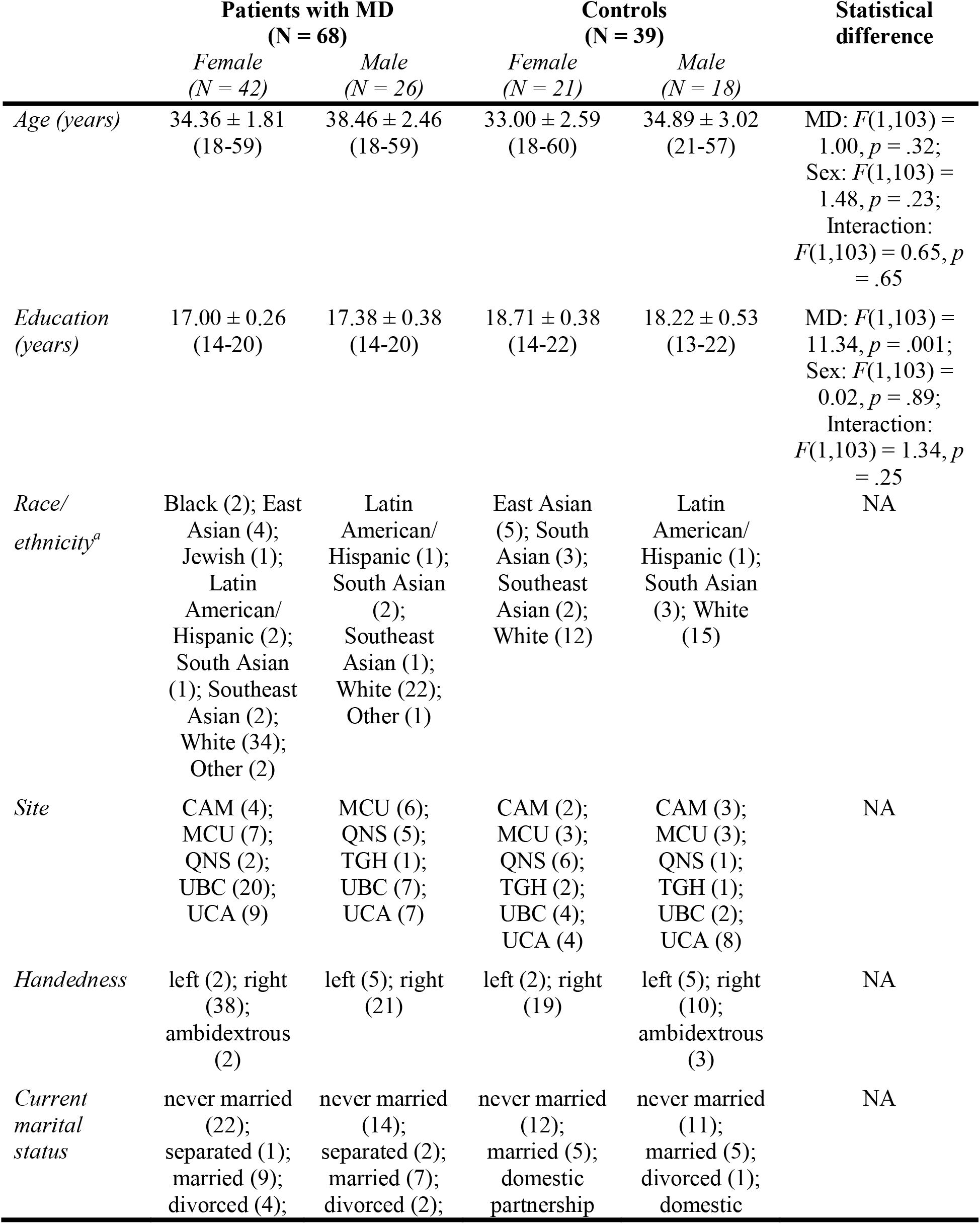
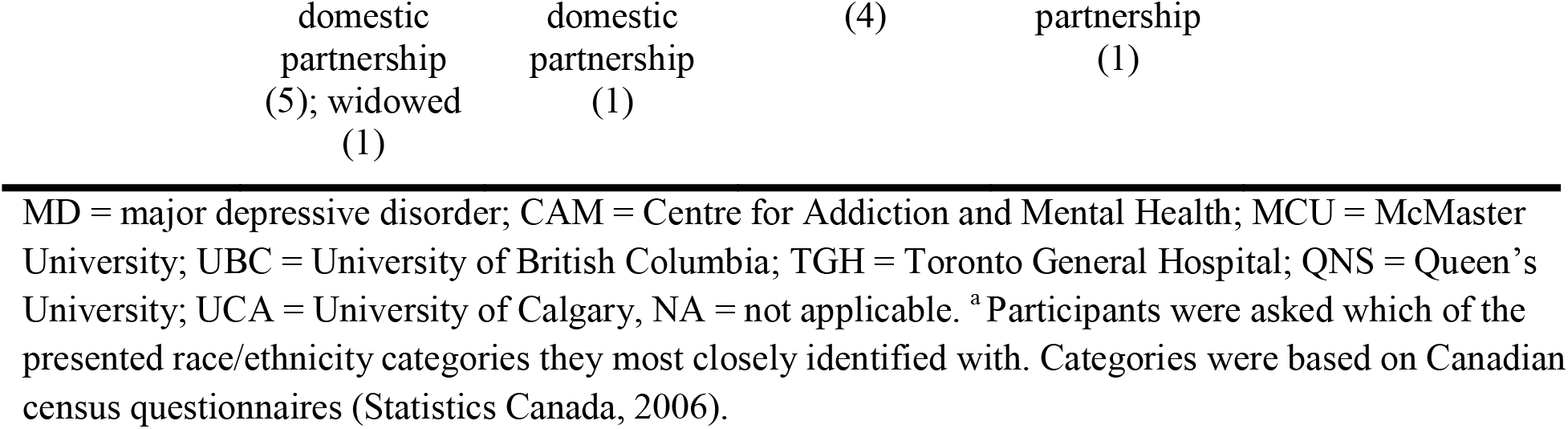
Demographic Characteristics of Female and Male Patients with MD and Controls (Mean ± SEM), and their Statistical Differences.

Diagnostic assessment using the Mini International Neuropsychiatric Interview (MINI; (Sheehan et al., 1998)) was performed by a trained research assistant to ensure MD participants met criteria for a major depressive episode according to the DSM-IV-TR, and entry level severity was confirmed using the Montgomery-Åsberg Depression Rating Scale (MADRS; (Montgomery & Åsberg, 1979)). All had MADRS scores >24 at enrollment, with their current episode lasting at least 3 months. Exclusion criteria for patients included meeting the diagnostic criteria for another psychiatric condition (except for anxiety), having a diagnosis of bipolar depression, experiencing psychotic symptoms in the current depressive episode, being at high risk for suicide or hypomanic switch, experiencing substance dependence in the last six months, being pregnant or breastfeeding, showing no response to four previous adequate pharmacotherapy interventions and having a previous unfavorable response to the medications used in the study. Patients on antidepressant medications prior to the study went through a wash-out period equivalent to five half-lives or more. Controls had no psychiatric diagnosis (as assessed with the MINI), and participants from both groups were excluded if they had a history of neurological conditions, head trauma or other unstable medical conditions, or any contraindications to MRI.

After the baseline visit was completed, patients received escitalopram at an initial dose of 10mg/d, increasing to 20mg/d after two or four weeks unless contra indicated by side effects. Every two weeks, patients’ symptom scores were assessed using the structured interview guide (SIGMA) for the MADRS (Williams & Kobak, 2008). Participants whose MADRS scores decreased >50% from baseline to week 8 were considered responders (N = 33), while participants whose MADRS scores decreased <50% (or increased) were considered non-responders (N = 35). Demographic information for both groups is presented in **Table 2**.

**Table 2.**
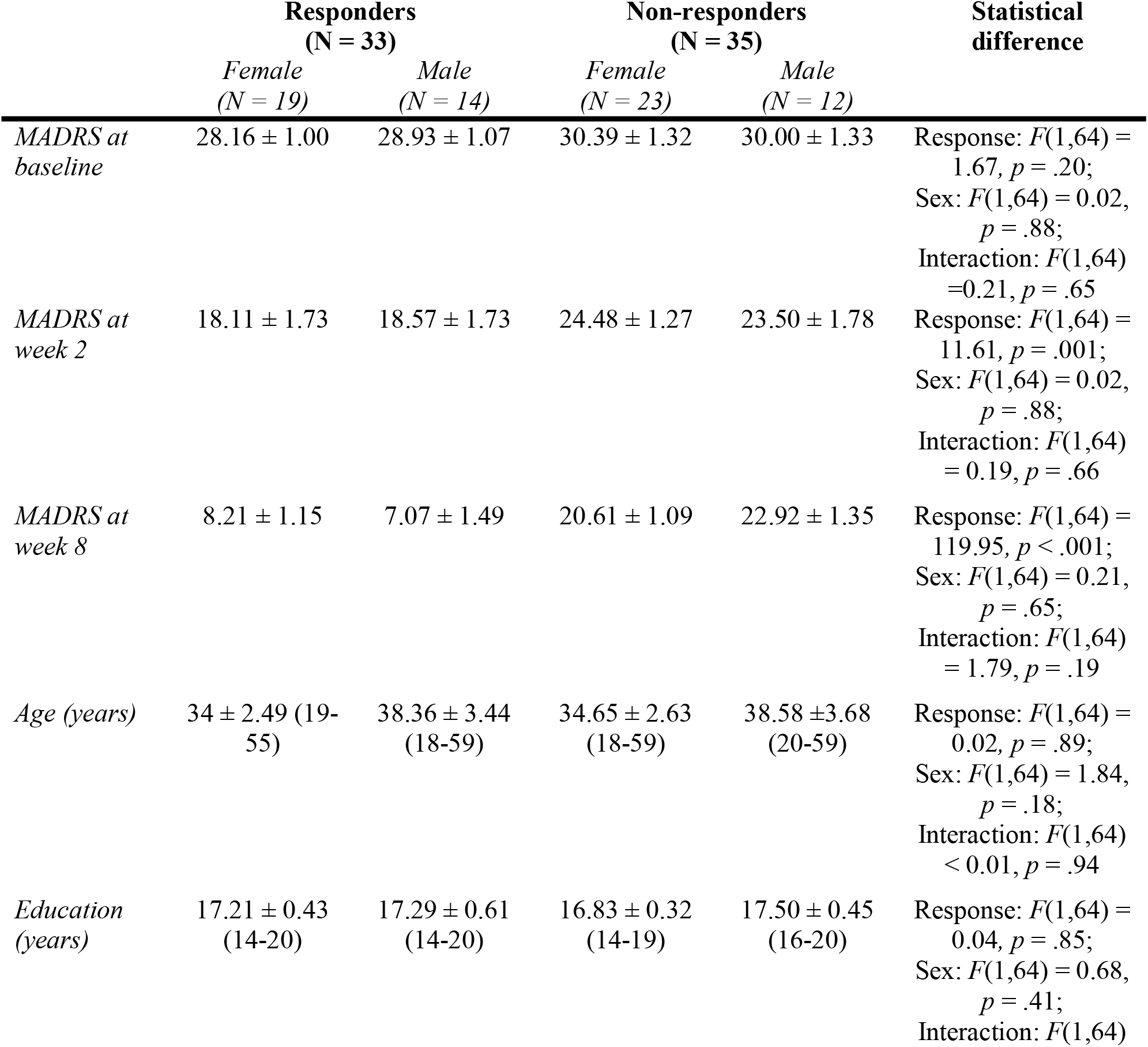
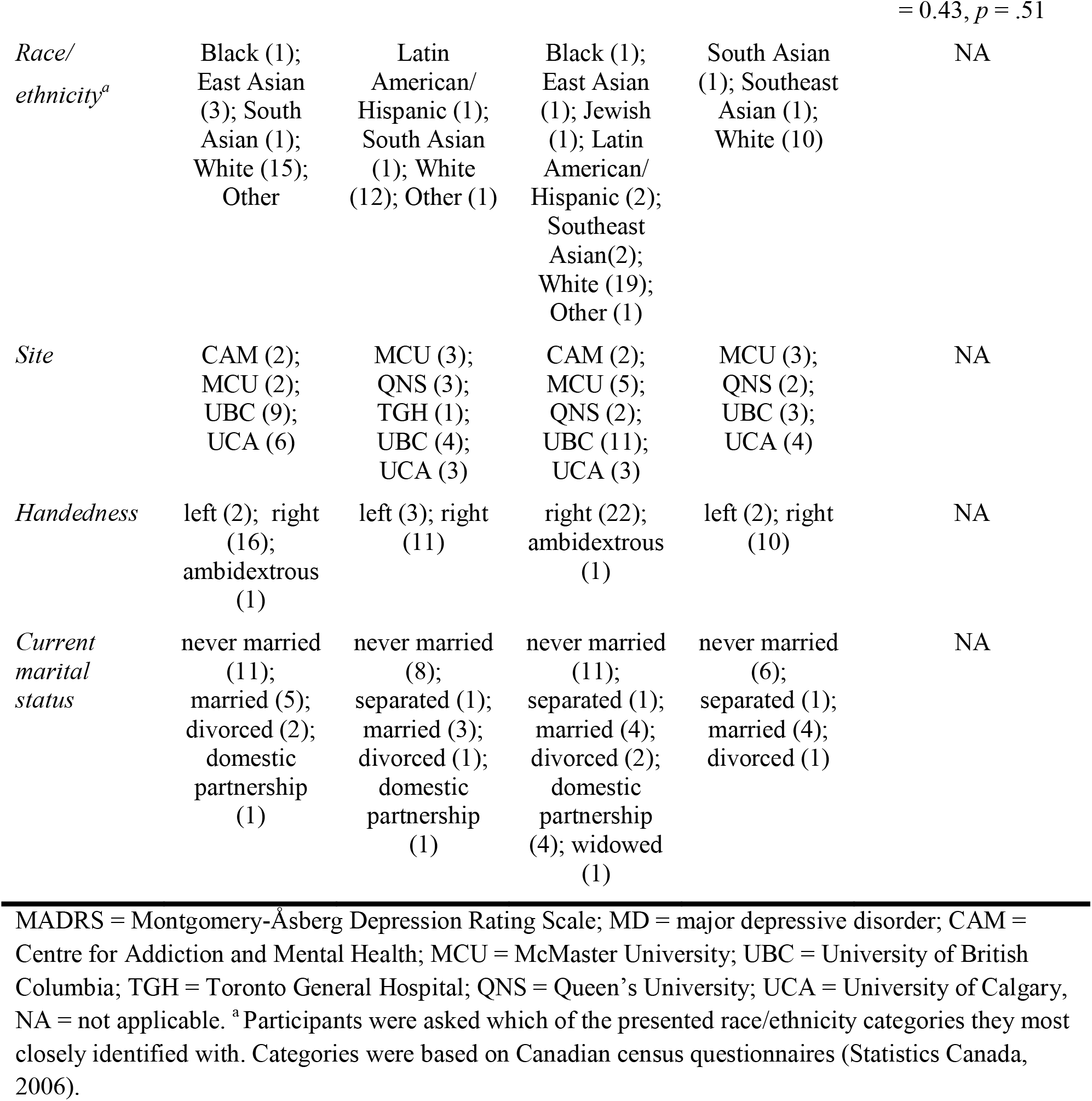
Clinical and Demographic Characteristics of Female and Male Responders and Non-Responders to Antidepressant Pharmacotherapy (Mean ± SEM), and their Statistical Differences.

|  | **Responders**<br>**(N = 33)** |  | **Non-responders**<br>**(N = 35)** |  | **Statistical**<br>**difference** |
| --- | --- | --- | --- | --- | --- |
|  | *Female*<br>*(N = 19)* | *Male*<br>*(N = 14)* | *Female*<br>*(N = 23)* | *Male*<br>*(N = 12)* |  |
| *MADRS at baseline* | 28.16 ± 1.00 | 28.93 ± 1.07 | 30.39 ± 1.32 | 30.00 ± 1.33 | Response: $F(1,64) = 1.67, p = .20$;<br>Sex: $F(1,64) = 0.02, p = .88$;<br>Interaction: $F(1,64) = 0.21, p = .65$ |
| *MADRS at week 2* | 18.11 ± 1.73 | 18.57 ± 1.73 | 24.48 ± 1.27 | 23.50 ± 1.78 | Response: $F(1,64) = 11.61, p = .001$;<br>Sex: $F(1,64) = 0.02, p = .88$;<br>Interaction: $F(1,64) = 0.19, p = .66$ |
| *MADRS at week 8* | 8.21 ± 1.15 | 7.07 ± 1.49 | 20.61 ± 1.09 | 22.92 ± 1.35 | Response: $F(1,64) = 119.95, p < .001$;<br>Sex: $F(1,64) = 0.21, p = .65$;<br>Interaction: $F(1,64) = 1.79, p = .19$ |
| *Age (years)* | 34 ± 2.49 (19-55) | 38.36 ± 3.44 (18-59) | 34.65 ± 2.63 (18-59) | 38.58 ± 3.68 (20-59) | Response: $F(1,64) = 0.02, p = .89$;<br>Sex: $F(1,64) = 1.84, p = .18$;<br>Interaction: $F(1,64) < 0.01, p = .94$ |
| *Education (years)* | 17.21 ± 0.43 (14-20) | 17.29 ± 0.61 (14-20) | 16.83 ± 0.32 (14-19) | 17.50 ± 0.45 (16-20) | Response: $F(1,64) = 0.04, p = .85$;<br>Sex: $F(1,64) = 0.68, p = .41$;<br>Interaction: $F(1,64)$ |
| = 0.43, $p = .51$ |  |  |  |  |  |
| *Race/ethnicity<sup>a</sup>* | Black (1); East Asian (3); South Asian (1); White (15); Other | Latin American/Hispanic (1); South Asian (1); White (12); Other (1) | Black (1); East Asian (1); Jewish (1); Latin American/Hispanic (2); Southeast Asian(2); White (19); Other (1) | South Asian (1); Southeast Asian (1); White (10) | NA |
| *Site* | CAM (2); MCU (2); UBC (9); UCA (6) | MCU (3); QNS (3); TGH (1); UBC (4); UCA (3) | CAM (2); MCU (5); QNS (2); UBC (11); UCA (3) | MCU (3); QNS (2); UBC (3); UCA (4) | NA |
| *Handedness* | left (2); right (16); ambidextrous (1) | left (3); right (11) | right (22); ambidextrous (1) | left (2); right (10) | NA |
| *Current marital status* | never married (11); married (5); divorced (2); domestic partnership (1) | never married (8); separated (1); married (3); divorced (1); domestic partnership (1) | never married (11); separated (1); married (4); divorced (2); domestic partnership (4); widowed (1) | never married (6); separated (1); married (4); divorced (1) | NA |

MADRS = Montgomery-Åsberg Depression Rating Scale; MD = major depressive disorder; CAM = Centre for Addiction and Mental Health; MCU = McMaster University; UBC = University of British Columbia; TGH = Toronto General Hospital; QNS = Queen's University; UCA = University of Calgary, NA = not applicable. <sup>a</sup>Participants were asked which of the presented race/ethnicity categories they most closely identified with. Categories were based on Canadian census questionnaires (Statistics Canada, 2006).

We completed power analyses prior to our planned analyses, using G*Power 3.1 (Faul et al., 2007). We estimated the effect size for the comparison of the common and individual effects based on results from Gratton et al. (Gratton et al., 2018) and found that our sample provides high statistical power (>0.99) for such effects. For effects that have not been investigated previously (e.g., sex, MD diagnosis and treatment response), we calculated statistical power for canonical small (Cohen’s *d* = 0.2), medium (*d* = 0.5) and large (*d* = 0.8) effect sizes, which revealed we had sufficient power to detect large and medium effects (power >0.86). This was sufficient for our study as we were interested in determining which sources of variance contributed the most, and were less interested in small effects.

### Tasks

During each scanning session, participants completed a 10-minute eyes-open resting state scan. They also performed a 10-minute affective go/no-go task, where they saw squares and circles on top of irrelevant emotional images (faces with angry or neutral expressions). They were instructed to press a button every time a circle appeared and inhibit their response when they saw a square. Stimuli were presented in a mixed block-event series design, with each block either showing only angry or only neutral faces. Participants completed 16 blocks in the same order. Lastly, they performed an 11.5-minute Monetary Incentive Delay (or anhedonia) task, during which they were instructed to press a button while a red square was presented on the screen. Before each target, a cue appeared indicating whether or not a reward would be given for a successful trial. Participants received feedback on each trial regarding response accuracy and reward status. These tasks have been previously described in (Lam et al., 2016; MacQueen et al., 2019). As we were interested in brain activity unrelated to the task, no data were excluded based on performance.

### fMRI data collection

Data were collected at the six sites using four different models of MRI scanners. All were 3.0 Tesla systems with multicoil phased-array head coils. Extensive quality control and standardization procedures were employed to ensure that data could validly be aggregated across scanner types and recording sites (Glover et al., 2012; MacQueen et al., 2019). The influence of variation across scanners on our analyses was also explored through supplementary analyses (see *Supplementary analyses* section below). Participants were scanned three times, at baseline, week 2 and week 8 of the study. Whole-brain T2*-sensitive blood oxygenation level dependent (BOLD) echo planar imaging (EPI) series was used to acquire the functional images with the following parameters: voxel dimensions (in mm) = 4×4×4, echo time (TE) = 25 or 30ms, repetition time (TR) = 2s, flip angle = 75° or 90°, field of view (FOV) = 256mm, matrix =64×64, number of slices = 34-40, acquisition order = interleaved (Lam et al., 2016; MacQueen et al., 2019).

### fMRI preprocessing

fMRI data were preprocessed with the OPPNI pipeline (Churchill et al., 2015, 2017; https://github.com/raamana/oppni) using the same parameters for resting state and task data. This pipeline involves motion correction, identification and interpolation of outliers, slice time correction, spatial smoothing across MRI scanners from different sites, regression of low frequency temporal trends, head motion estimates, global signal modulations and physiological noise, low pass filtering and spatial normalization to a structural MNI template. In addition, the data were directly registered to a sample-specific EPI template through an affine transformation followed by a nonlinear transformation following the EPInorm strategy (Calhoun et al., 2017). A more detailed description of these steps can be found in (van der Wijk et al., 2021), which used the same parameters.

Next, we regressed out task-evoked activity from the affective go/no-go and anhedonia data for each participant and session using finite impulse response (FIR) task regression, which provides a flexible fit in terms of the exact shape of the HRF for each individual (Cole et al., 2019). This and the following steps were completed in MATLAB 2018b (The MathWorks, Inc., Natick, Massachusetts). We used code provided by Cole et al. ((Cole et al., 2019); firLag parameter = 10 TRs/20 seconds) to create time-locked FIR regressors from the following primary regressors. For the affective go/no-go task, we used one for the instruction slides, and one each for the four types of blocks. For the anhedonia task, we used two for the cues, one for the target, and four for each type of feedback. An intercept was also added to each regression model. The task-residualized data were used for further analysis. Next, we subtracted the mean of the time course for each voxel from each data point for that voxel. Last, we concatenated the resting state and task data to create time courses of ∼30 minutes per participant and session.

### Parcellation

We parcellated each individual’s brain into 333-regions (Gordon et al., 2016). Seven regions (six in the orbitofrontal cortex and one in the right inferior temporal gyrus) did not have coverage and were therefore excluded. The average time course over the voxels within each region was extracted and used for the functional connectivity estimation.

### Data analysis

We performed analyses on controls-only, patients and controls together, and patients-only to examine the contribution of different sources of variance following the methodology in (Gratton et al., 2018). First, we estimated functional connectivity between all regions within each individual and session using product-moment correlations between the ROI time courses and applied a Fisher’s z-transformation. Next, we correlated the upper half of each session and individual’s whole-brain connectivity matrix with that of each other session (including sessions from the same individual) and individual’s whole-brain connectivity pattern to construct a similarity matrix (see **Figure 1c**). In this similarity matrix, each row and column represented a specific individual and session, together depicting the similarity of whole-brain functional connectivity for different pairs of individuals and sessions. We applied Fisher’s z-transformation to these similarity matrices as well.

**Figure 1.**
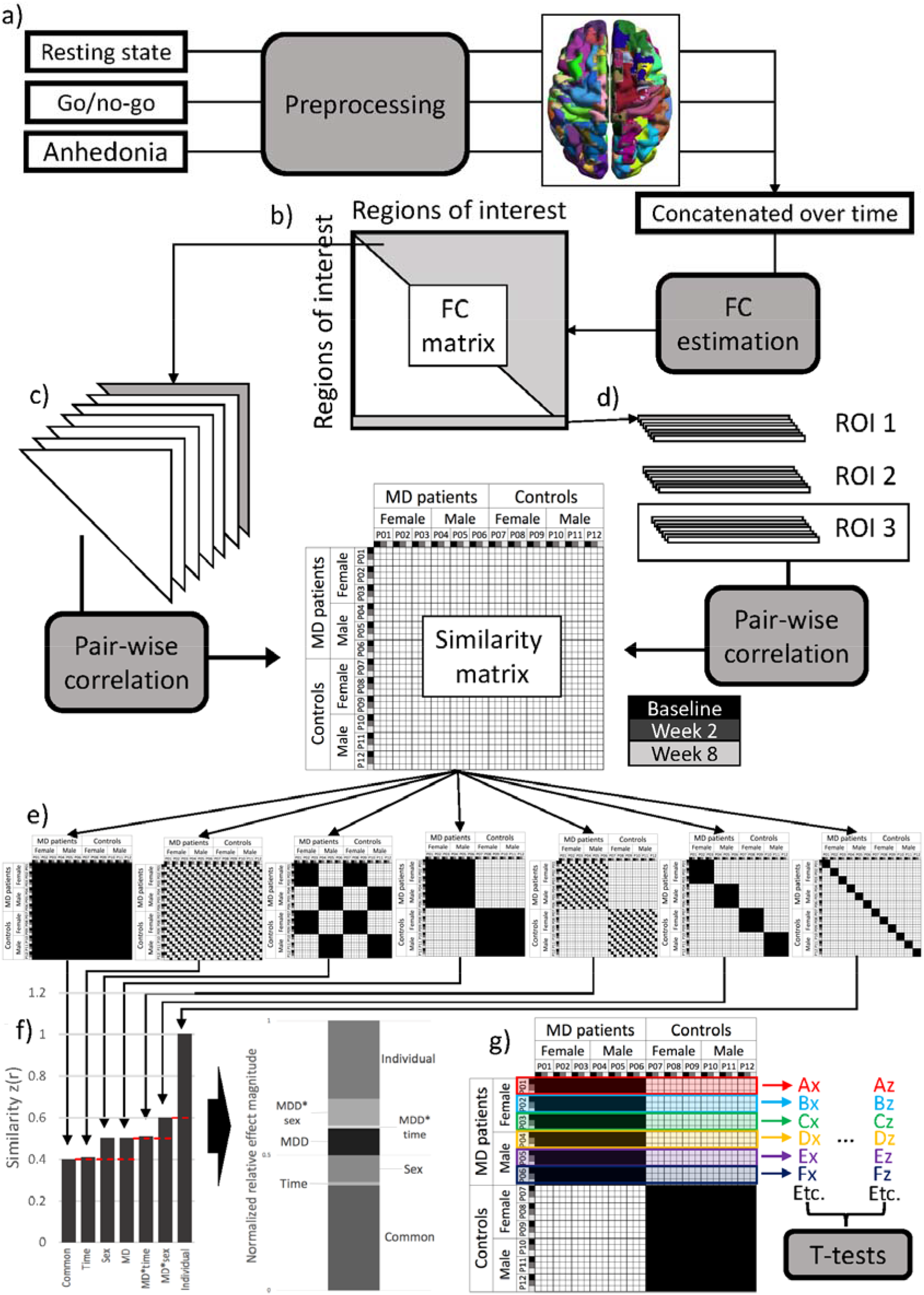
Overview of analysis procedure. The analysis for patients and controls is used as an example, but steps were the same for analyses with controls only and patients only. a) The fMRI data from resting state, affective go/no-go and anhedonia (or monetary incentive delay) tasks were preprocessed using the OPPNI pipeline (Churchill et al., 2015, 2017), parcellated into 326 regions and concatenated for each participant and session. b) FC was calculated as the correlation between the time courses of each region pair, resulting in a region-by-region FC matrix. c) For the whole-brain analyses, the upper triangle (grey) of the FC matrix from each participant and session was correlated with that of every other participant and session to create a similarity matrix. d) For the region-specific analyses, a similarity matrix was constructed for each ROI by correlating each ROI’s row in the correlation matrix with that of the same row of other participants and sessions. e) The contribution of each source of variance was estimated by calculating the average similarity over different configurations of the similarity matrix. From left to right, the presented patterns display the configuration for the calculation of the common effect (similar FC across all participants and sessions), the session effect (similar FC across participants within sessions), the sex effect (similar FC among female and among male participants), the MD effect (similar FC among patients with an MD diagnosis and among controls), the MD*session interaction (similar FC among patients and controls within sessions), the MD*sex interaction (similar FC among female patients, male patients, female controls and male controls), and the individual effect (similar FC within individuals across sessions). f) From the average similarity for each effect, we calculated the normalized relative effect magnitude by first subtracting the baseline for each effect (indicated by the dashed red lines) and then dividing by the total relative similarity. For the region-specific analyses, we visualized the normalized relative effect magnitudes but did not perform statistical comparisons. g) For the whole-brain analyses, we also calculated the average similarity for each effect per individual by taking the average of the patterns for the three rows representing each participant. The example shows this for the MD effect. For each participant, indicated by different color outlines, we calculated the average similarity of that participant to all other participants and sessions within the same group (i.e., by taking the average across black squares within the three rows representing the three recording sessions of that participant). This way, we calculated each of the effects shown in part e of the figure at an individual level as well. These participant-specific average similarity values were then entered into dependent samples t-tests to examine differences in effect magnitudes. Each capital letters (A-F) represent individuals and the lowercase letters (x-z) the different effects (i.e., Ax is the MD effect magnitude for participant 1, Az is the effect magnitude for a different effect for that same participant). FC = functional connectivity; ROI = region of interest; MD = major depressive disorder

To quantify the magnitude of different sources of variance, we calculated the average over different parts of the similarity matrix (as illustrated in **Figure 1e**). The diagonal of the matrix (all ones by definition) was not included in the averages. Each average was also calculated within the three rows representing each participant to get an estimate of each effect based on the similarity values of that participant with their own and the other participants’ data (see **Figure 1g**). These participant-specific values were submitted to dependent sample t-tests in MATLAB with permutation testing (1000 permutations) to estimate significance. While the data were not fully independent, they were exchangeable, which is what is required for permutation testing (LaFleur & Greevy, 2009).

We compared each effect to its baseline (see **Figure 1f**) and the other effects of the same type (e.g., main effects to other main effects), meaning we conducted four statistical comparisons for the controls-only, and ten comparisons for each of the patients and controls and patients-only samples. We used the false-discovery rate (FDR) method to correct for multiple comparisons (Benjamini & Yekutieli, 2001). We calculated the normalized relative effect magnitude as an indication of effect size. First, we subtracted the baseline from each effect (see **Figure 1f**), and then divided by the sum of all baseline-corrected magnitudes. As such, normalized relative effect magnitude values reflect the proportion of each effect that is unique, i.e., not explained by its baseline, relative to the combined unique magnitude of all effects. For the interaction effects, we used the main effect with the highest similarity as baseline (e.g., if the MD effect was larger than the sex effect, the MD effect would serve as the baseline for the MD*sex interaction). If an effect had a lower magnitude than its baseline, the baseline-corrected magnitude was set to zero.

We also examined the distribution of these effects in the brain by conducting the same analysis for each region separately, i.e., instead of correlating the upper-triangle of the whole-brain connectivity matrix, we correlated the connectivity estimates of one brain region with all other brain regions across participants and sessions to create a similarity matrix for each region (see **Figure 1d**). We calculated the normalized relative effect magnitude for each effect and region to illustrate where in the brain the effects were most prominent, but did not perform statistical analyses on these region-specific estimates.

### Supplementary analyses

We conducted several supplementary analyses to ensure the validity of our main analyses. First, we performed the whole-brain analyses excluding ROIs with fewer than eight voxels (45/326 ROIs excluded), as the average time courses extracted from these ROIs may have been less reliable/accurate. Next, we conducted the whole-brain analyses for patients and controls, and patients only on scanner, sex, age and education matched subsamples to examine if these factors may have influenced our results. In addition, these analyses served to ascertain if unequal group sizes may have distorted our findings. More methodological details (e.g., about the matching procedure) and the results of these analyses are reported in the supplementary materials.

### Code & data accessibility

All custom codes for these analyses are available at https://osf.io/79gv8/files/github?view_only=9e105962ce4c4ebf8cf35393471a7b69. The data used in this manuscript has been collected as part of the CAN-BIND initiative, an Integrated Discovery Program of the Ontario Brain Institute (OBI). OBI has released data from CAN-BIND’s foundational study which aims to identify biomarkers that predict treatment response in people with depression. The dataset currently available on Brain-CODE includes baseline and longitudinal data from participant follow-up visits in weeks 2 to 8 of the study (Phase 1). All data have been standardized, cleaned and curated to maximize utility for analysis across different data modalities, and imaging data was converted to a BIDS-friendly naming convention. For access requirements, please visit OBI’s Brain-CODE Neuroinformatics Platform (https://www.braincode.ca/) or.

## Results

### Participants

The demographic information for patients and controls, broken into female and male participant subgroups, is presented in **Table 1**. Two 2 (patients vs controls) x2 (female vs male participants) ANOVAs indicated there was no significant effect of age (all *F*s < 1.48, all *p*s > .23). However, there was a significant main effect of MD diagnosis for years of education (*F*(1,103) = 11.34, p = .001). Namely, participants in the control group (*M* = 18.49, *SD* = 1.97) had a higher number of years of education compared to the participants in the patient group (*M* = 17.15, *SD* = 1.79), which is consistent with previous studies alike. We examined the potential impact of this difference in our supplementary analyses and found no evidence of an effect (see *Supplementary materials*). **Table 2** displays the clinical and demographic information for responder and non-responder groups, again broken down into female and male participant subgroups. As expected, there was a significant main effect of response for MADRS symptom scores at week 2 (*F*(1,64) = 11.61, *p* = .001) and week 8 (*F*(1,64) = 119.95, *p* < .001). Apart from that, no significant effects were observed.

### Sources of variance in whole-brain FC

We investigated the contributions of different sources of variance relative to their baselines in controls-only, patients and controls together and patients-only. The pattern was similar across all three samples (see **Figure 2** and **Table 3)**. Namely, common FC across all participants and conditions (common effect) and individual-specific FC (individual effect) contributed most to the observed variance (normalized relative effect magnitude = 50.9-51.5% and 46.0-47.5% respectively). The individual effect was significantly bigger than its baseline in all samples (*p*s < .001). The contributions of sex, MD, response effects and their interactions were significant but small (normalized relative effect magnitude = 0.3-1.2%, *p*s < .001). The session effect and its interactions were significantly smaller than their baselines (*p*s < .001).

**Figure 2.**
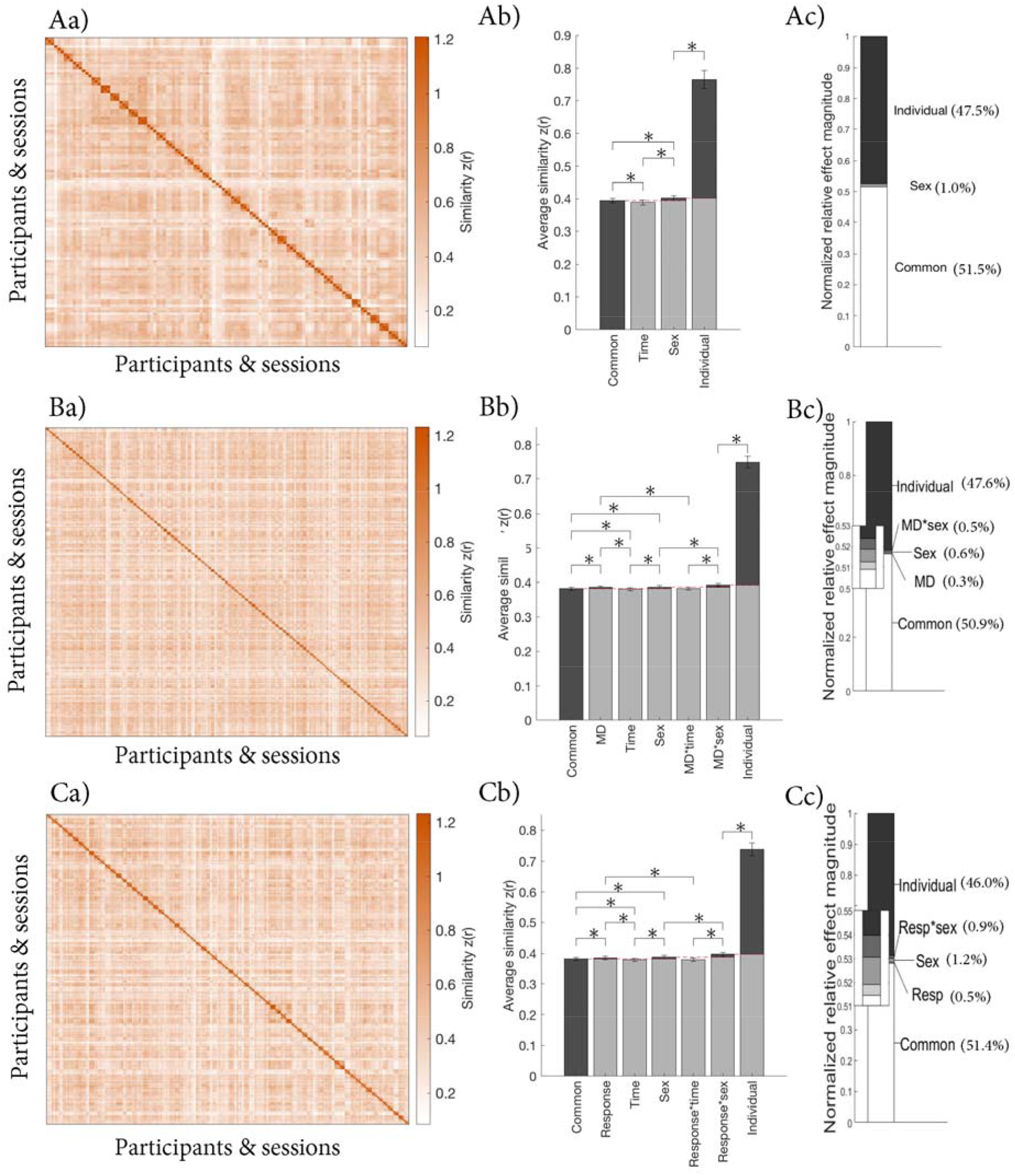
Contributions of different sources of variance in FC in A) controls-only, B) patients and controls, and C) patients-only. a) Similarity matrix displaying the Fisher transformed correlations between the whole-brain FC of different participants and sessions. The matrices are organized by group (sex and MD diagnosis or response), individual and session (baseline, week 2 and week 8) as illustrated in **Figure 1**; b) Average similarity for each effect. **Figure 1e** illustrates how each average was calculated. The red dashed lines indicate the baseline for each effect, which was subtracted to calculate the normalized relative effect magnitudes; c) Normalized relative effect magnitude for each effect.

**Table 3.**
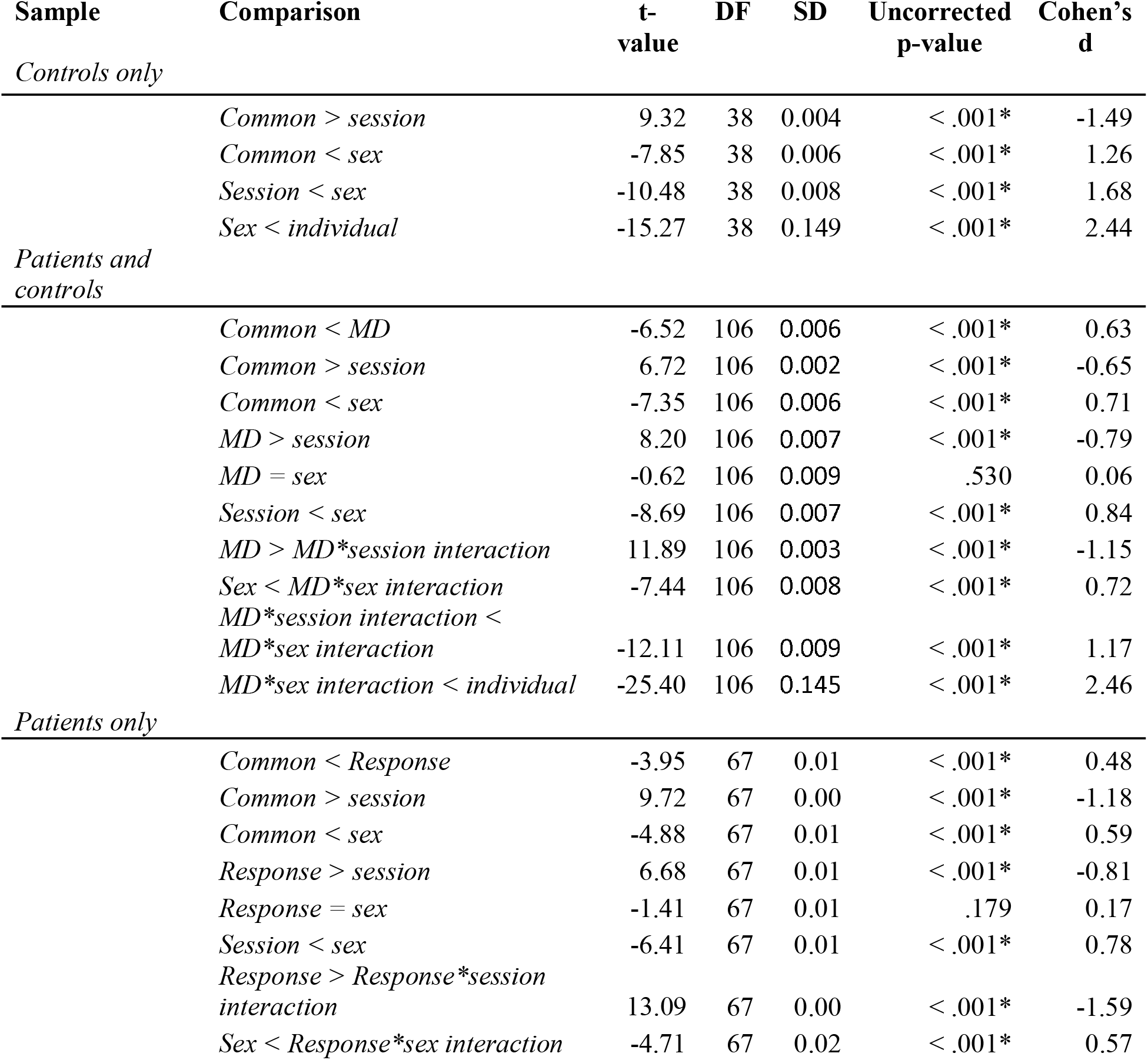
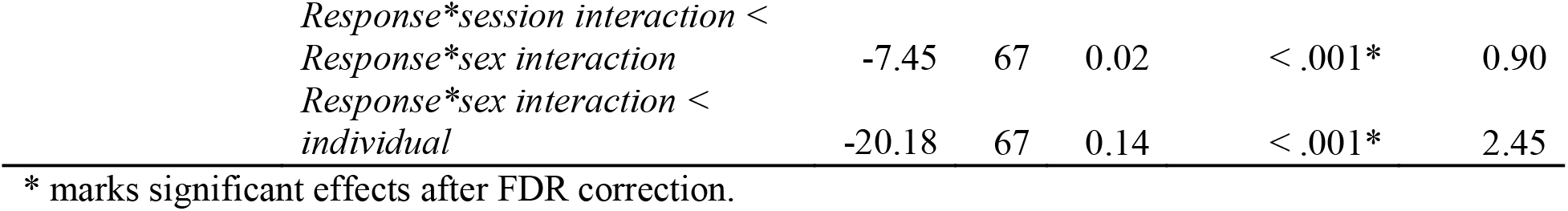
Results of Paired Samples T-Tests to Compare Magnitude of Effects in Controls Only, Patients and Controls and Patients Only.

### Localization of effects

We also illustrate the normalized relative effect magnitudes calculated per ROI to indicate where each effect was most prominently present in the brain. As the distributions of the common and individual effects were similar across samples, we present the results from the patients and controls together in **Figure 3** (patients-only results can be found in **Figure S1**). In both samples, the common effect was most strongly present in default mode network, somatosensory, motor, visual and auditory areas. The individual effect was expressed most strongly in frontoparietal, dorsal attention and cingulo-parietal network areas. The sex, MD, and response effects and their interactions contributed relatively little, therefore the distribution of these effects cannot be seen on the same scale as the common and individual effects. However, we illustrate them on a narrower scale in **Figure S2.**

**Figure 3.**
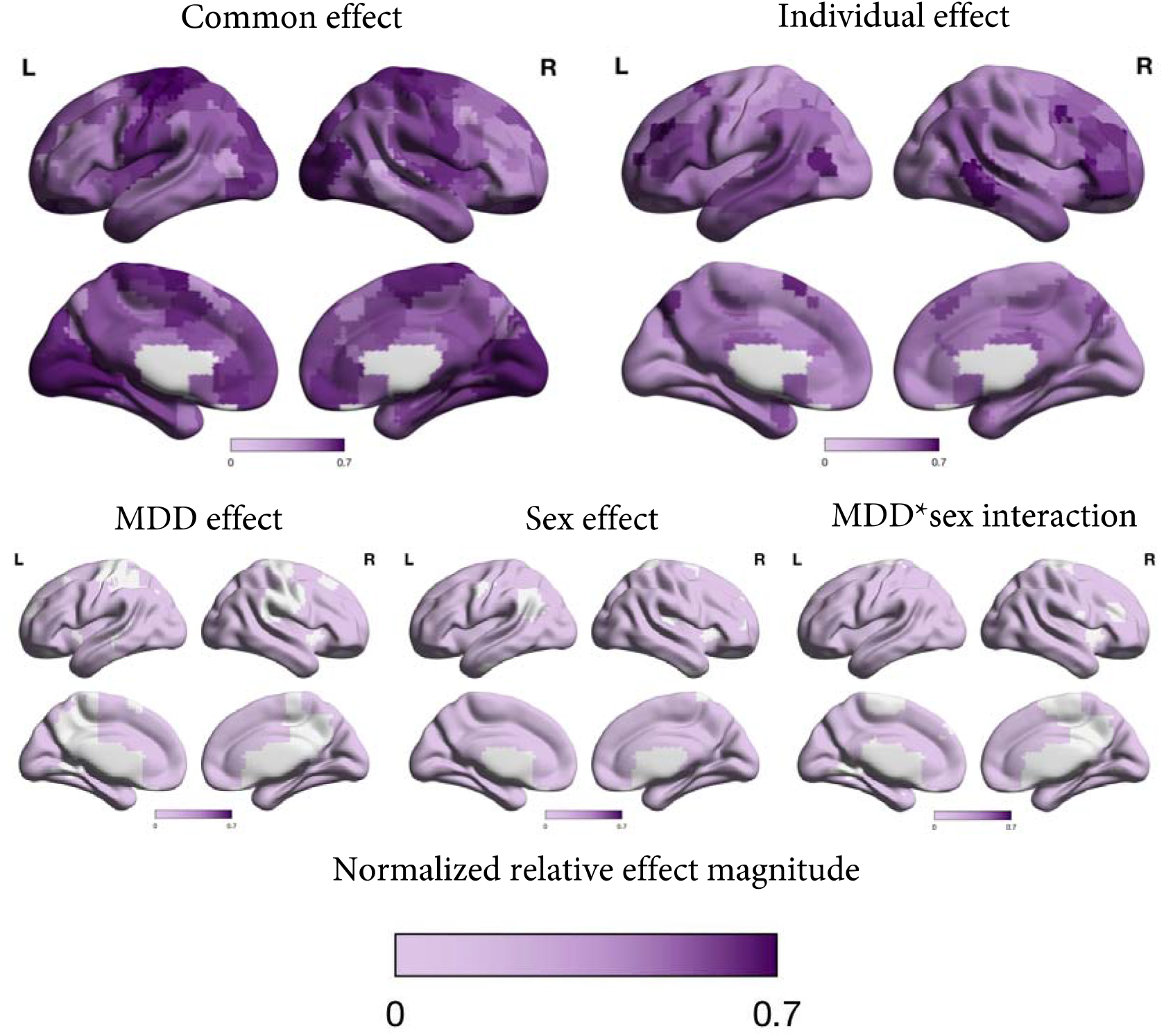
Normalized relative effect magnitude in each region of the brain for each effect (common, MD, Sex, MD*sex and individual) in patients and controls. The MD, sex and MD*sex effects are presented on a narrower colour scale in **Figure S2** so the localization of these effects can be distinguished.

### Exploratory analyses

As the results above point to a large contribution of individual variation relative to other sources of variance (e.g., MD diagnosis, sex and response to treatment), we further explored the contributions of individual*task and individual*session interactions (see **Figure S5**) in exploratory analyses. Instead of pooling data across tasks, we calculated functional connectivity for each task and session. We had 9.8 minutes of data for the resting state and affective go/no-go task, and 11.5 minutes for the anhedonia task, which provide ∼72-82% reliability for the individual connectivity matrices (Gordon et al., 2017; Laumann et al., 2015).

These exploratory analyses revealed a similar pattern of results for the effects examined with the pooled data (see **Table 4 and Figure 4**). The only notable difference was that the session effect was no longer smaller than its baseline for controls and patients, and for patients-only. In addition, we found a significant contribution of the individual*task interaction for all three samples (normalized relative effect magnitude = 14.2-15.0%, all *p*s < .001). Interestingly, the individual*session interaction was significant in patients-only (normalized relative effect magnitude = 2.1%, *p* = .016), but not in the other two samples.

**Figure 4.**
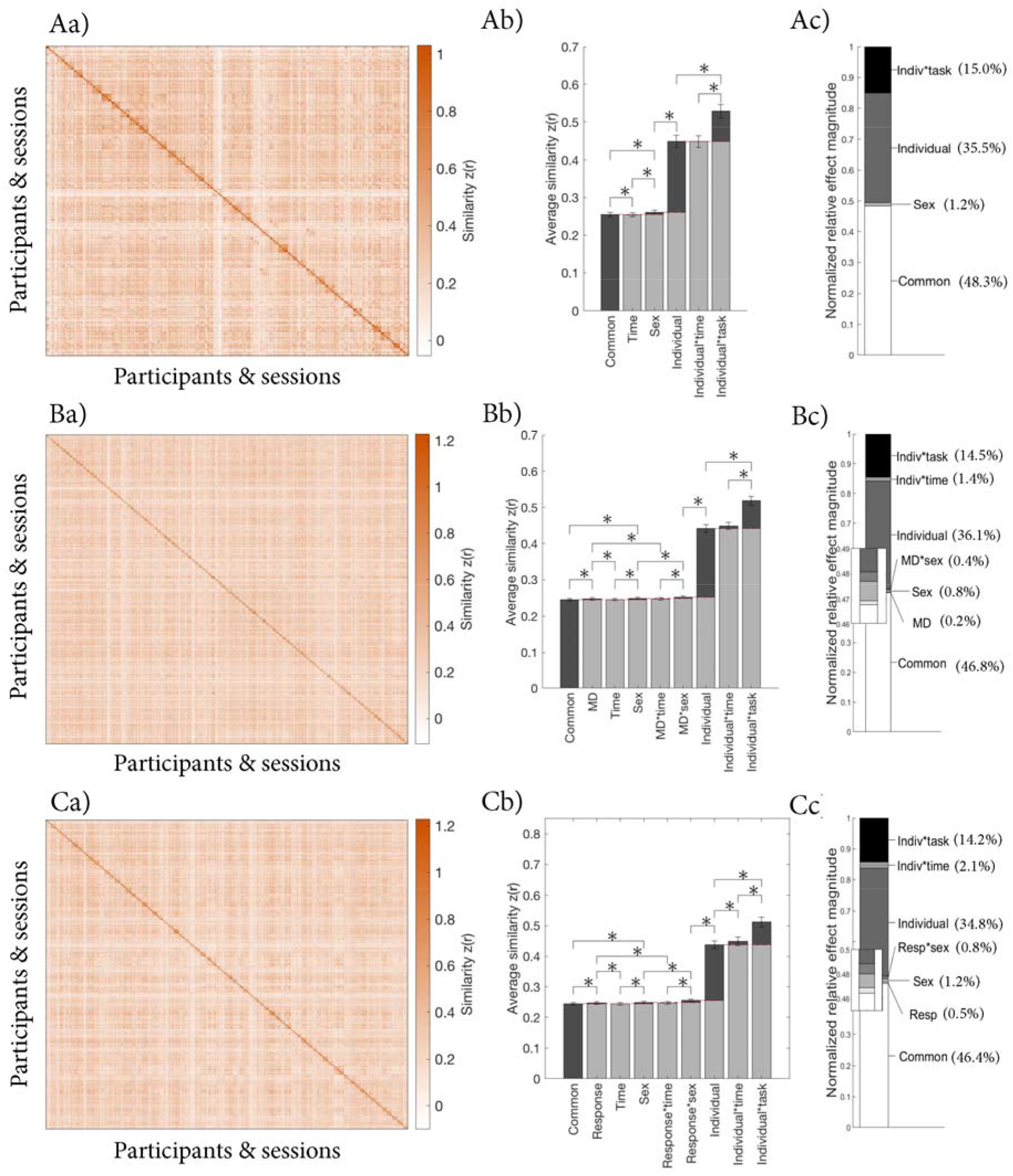
Contributions of different sources of variance in FC as examined in our exploratory analyses in A) controls only, B) patients and controls, and C) patients only. a) Similarity matrix displaying the Fisher transformed correlations between the whole-brain FC of different participants and sessions. The matrices are organized by group (sex and MD diagnosis or response), individual, task (resting state, affective go/no-go and anhedonia) and session (baseline, week 2 and week 8) as illustrated in **Figure S5**; b) Average similarity for each effect. See **Figure S5** for the pattern across which each average was calculated. The red dashed lines indicate the baseline for each effect, which were subtracted to calculate the normalized relative effect magnitudes; c) Normalized relative effect magnitude for each effect.

**Table 4.**
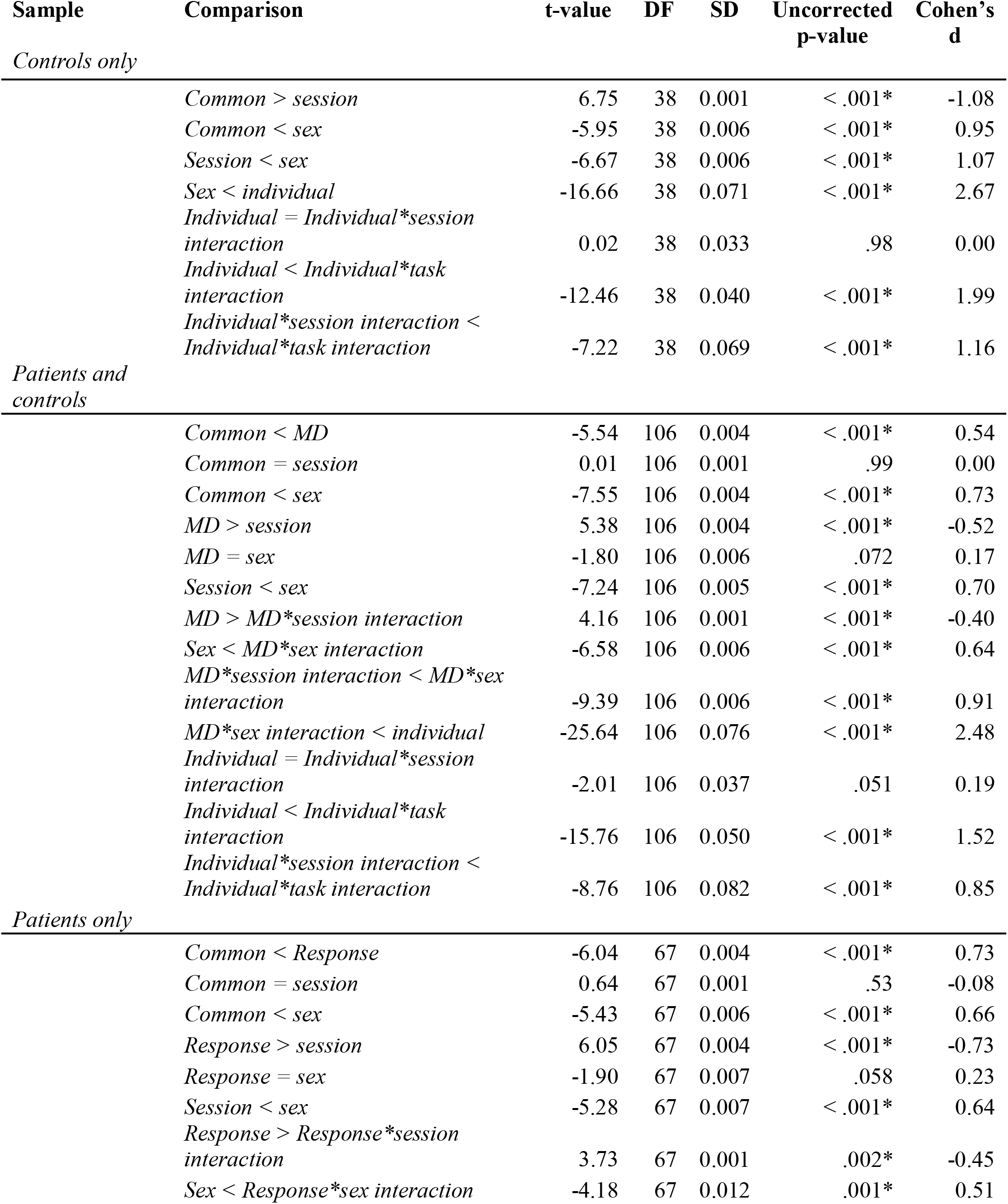
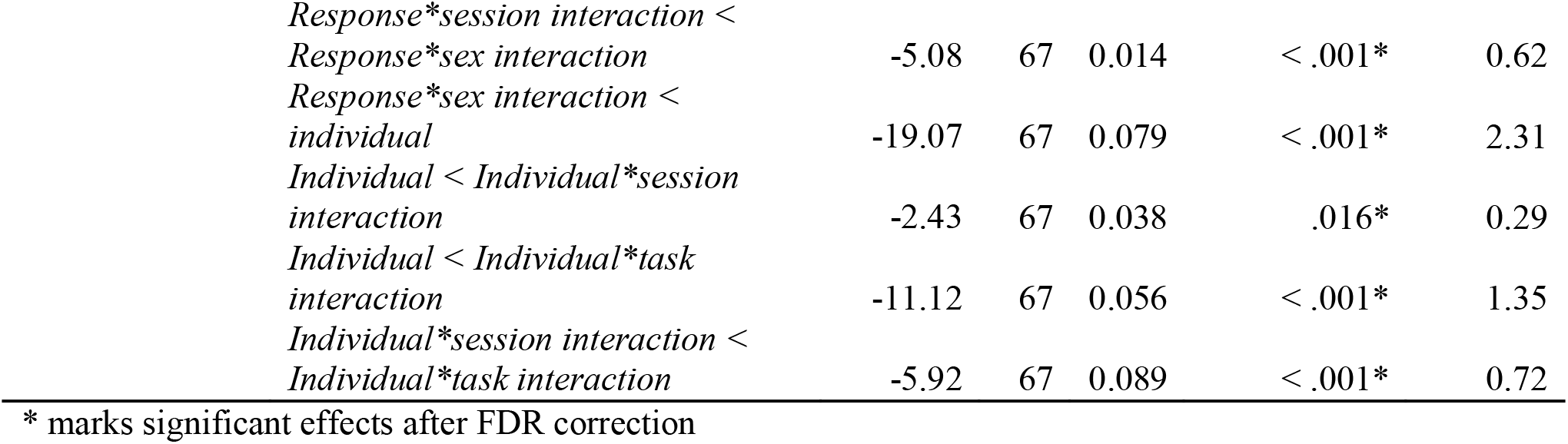
Results of Paired Samples T-Tests to Compare Magnitude of Effects in Our Exploratory Analyses in Controls Only, Patients and Controls and Patients Only.

| Sample | Comparison | t-value | DF | SD | Uncorrected p-value | Cohen's d |
| --- | --- | --- | --- | --- | --- | --- |
| *Controls only* |  |  |  |  |  |  |
|  | *Common > session* | 6.75 | 38 | 0.001 | < .001* | -1.08 |
|  | *Common < sex* | -5.95 | 38 | 0.006 | < .001* | 0.95 |
|  | *Session < sex* | -6.67 | 38 | 0.006 | < .001* | 1.07 |
|  | *Sex < individual* | -16.66 | 38 | 0.071 | < .001* | 2.67 |
|  | *Individual = Individual\*session interaction* | 0.02 | 38 | 0.033 | .98 | 0.00 |
|  | *Individual < Individual\*task interaction* | -12.46 | 38 | 0.040 | < .001* | 1.99 |
|  | *Individual\*session interaction < Individual\*task interaction* | -7.22 | 38 | 0.069 | < .001* | 1.16 |
| *Patients and controls* |  |  |  |  |  |  |
|  | *Common < MD* | -5.54 | 106 | 0.004 | < .001* | 0.54 |
|  | *Common = session* | 0.01 | 106 | 0.001 | .99 | 0.00 |
|  | *Common < sex* | -7.55 | 106 | 0.004 | < .001* | 0.73 |
|  | *MD > session* | 5.38 | 106 | 0.004 | < .001* | -0.52 |
|  | *MD = sex* | -1.80 | 106 | 0.006 | .072 | 0.17 |
|  | *Session < sex* | -7.24 | 106 | 0.005 | < .001* | 0.70 |
|  | *MD > MD\*session interaction* | 4.16 | 106 | 0.001 | < .001* | -0.40 |
|  | *Sex < MD\*sex interaction* | -6.58 | 106 | 0.006 | < .001* | 0.64 |
|  | *MD\*session interaction < MD\*sex interaction* | -9.39 | 106 | 0.006 | < .001* | 0.91 |
|  | *MD\*sex interaction < individual* | -25.64 | 106 | 0.076 | < .001* | 2.48 |
|  | *Individual = Individual\*session interaction* | -2.01 | 106 | 0.037 | .051 | 0.19 |
|  | *Individual < Individual\*task interaction* | -15.76 | 106 | 0.050 | < .001* | 1.52 |
|  | *Individual\*session interaction < Individual\*task interaction* | -8.76 | 106 | 0.082 | < .001* | 0.85 |
| *Patients only* |  |  |  |  |  |  |
|  | *Common < Response* | -6.04 | 67 | 0.004 | < .001* | 0.73 |
|  | *Common = session* | 0.64 | 67 | 0.001 | .53 | -0.08 |
|  | *Common < sex* | -5.43 | 67 | 0.006 | < .001* | 0.66 |
|  | *Response > session* | 6.05 | 67 | 0.004 | < .001* | -0.73 |
|  | *Response = sex* | -1.90 | 67 | 0.007 | .058 | 0.23 |
|  | *Session < sex* | -5.28 | 67 | 0.007 | < .001* | 0.64 |
|  | *Response > Response\*session interaction* | 3.73 | 67 | 0.001 | .002* | -0.45 |
|  | *Sex < Response\*sex interaction* | -4.18 | 67 | 0.012 | .001* | 0.51 |

|  |  |  |  |  |  |
| --- | --- | --- | --- | --- | --- |
| *Response\*session interaction < Response\*sex interaction* | -5.08 | 67 | 0.014 | < .001* | 0.62 |
| *Response\*sex interaction < individual* | -19.07 | 67 | 0.079 | < .001* | 2.31 |
| *Individual < Individual\*session interaction* | -2.43 | 67 | 0.038 | .016* | 0.29 |
| *Individual < Individual\*task interaction* | -11.12 | 67 | 0.056 | < .001* | 1.35 |
| *Individual\*session interaction < Individual\*task interaction* | -5.92 | 67 | 0.089 | < .001* | 0.72 |

\* marks significant effects after FDR correction

We also examined regional localization for these effects. We illustrate these for patients-only in **Figure 5** (patients and controls results can be found in **Figure 6)**. The individual*task interaction was most prominent in right lateral prefrontal cortex, bilateral visual areas, and left somatosensory areas. The individual*session interaction was expressed primarily in bilateral subgenual cingulate cortex, medial temporal lobes and temporal poles.

**Figure 5.**
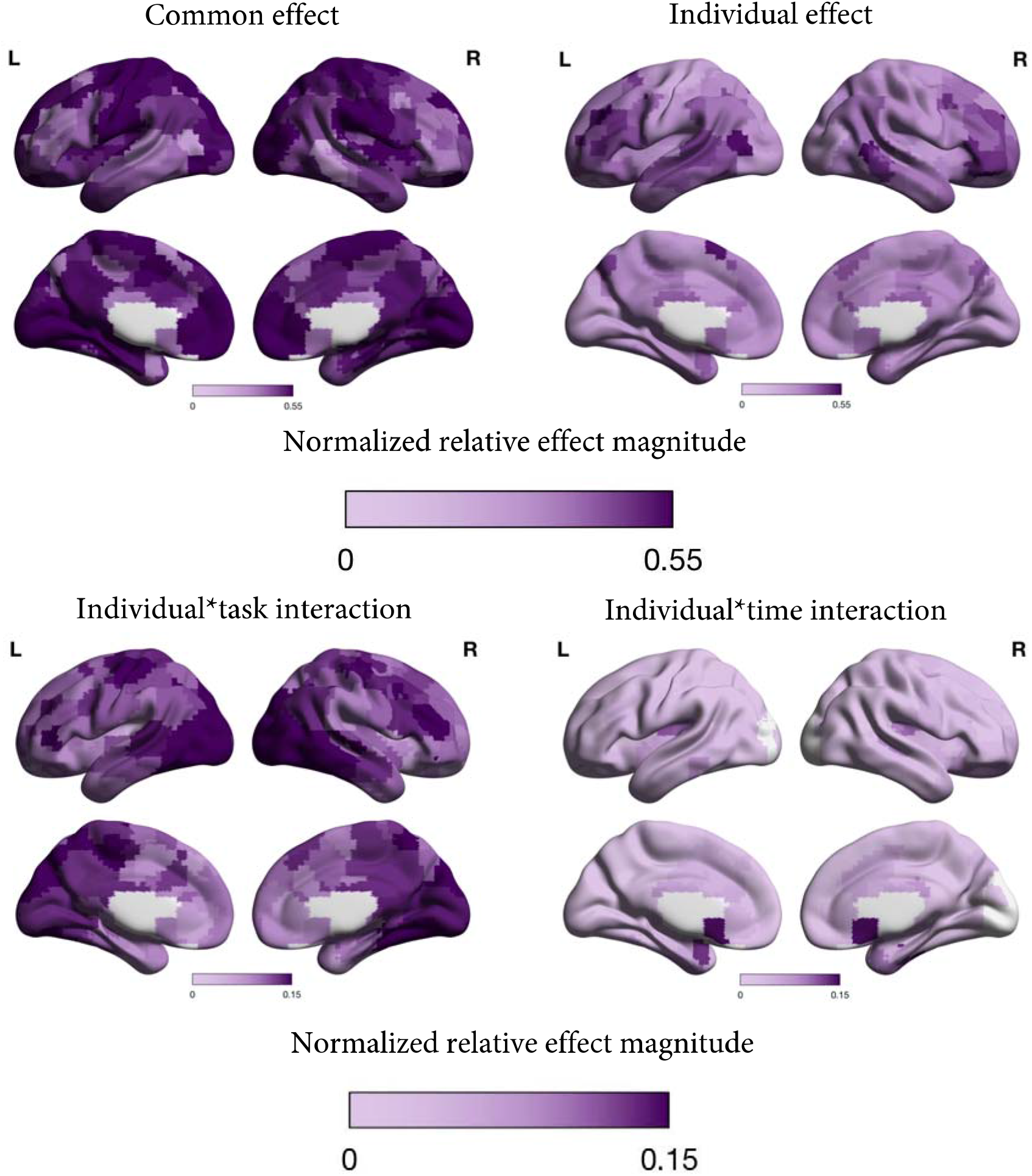
Normalized relative effect magnitude in each region of the brain for selected effects from the exploratory analyses (common, individual, individual*task, and individual*session) in patients only.

## Discussion

In this study, we investigated the relative contributions of several sources of variance observed in FC data collected across 8 weeks from participants with MD and controls, with a particular interest in the relative contribution of clinical characteristics including MD diagnosis and response to treatment. While commonly studied sources (MD diagnosis, response to/change with treatment and biological sex) did contribute to the overall variance, these effects were small (0.3-1.2%) relative to common and individual-specific FC, which explained >95% of the variance in the data across analyses. Supplementary analyses showed that ROI size, scanner types, demographics or unequal group sizes did not meaningfully influence our findings. The localization analyses showed brain areas with low and high individual variation in line with previous findings in neurotypical individuals.

The pattern of relative contributions of common versus individual variance in FC in our study resembled that reported by Gratton et al. (2018) in a small (N = 10) highly sampled group of neurotypical individuals (i.e., that common connectivity across all sessions and participants and individual differences contributed most to the overall variance). The regions where individual variation was low (strong common FC) or high (strong individual-specific FC) further matched previous work with neurotypical adults (Chen et al., 2015; Gratton et al., 2018; Kong et al., 2019; Mueller et al., 2013; Seitzman et al., 2019). Namely, individual variation was low in primary sensory and motor, as well as some default mode network areas, and high in lateral (pre)frontal areas and mid temporal areas. Whereas sensorimotor networks develop early in life and are therefore more strongly guided by genetic determination (but also see Graff et al., 2022), association networks continue to develop long after birth and may therefore be more impacted by environmental factors, leading to greater inter-individual variation (Mueller et al., 2013). These findings indicate a degree of stability in the distribution and magnitude of shared and individual variation in FC regardless of mental health and treatment status.

To our knowledge, this study is the first to examine the relative magnitudes of effects related to MD diagnosis, response to treatment and biological sex. The results suggest that these effects were small, which is consistent with studies revealing significant but small group differences next to a great degree of overlap in structural and functional brain features across groups (Joel et al., 2015; Winter et al., 2021). Considered together with large commonalities and individual differences, the small relative magnitude of these group effects may contribute significantly to the variable findings in neuroimaging features of MD and treatment response to date. Further, these findings may also shed some light on why, despite hopes to the contrary, it is proving difficult to use such features to predict treatment success at an individual level (Etkin, 2019; Gratton et al., 2020). As explained by Arbabshirani et al. (2017), large overlap between groups generally results in low classification rates, even when mean group differences are highly significant. Next to discriminatory value, ensuring that individual estimates are reliable (e.g., through the collection of greater amounts of data per individual) will likely be essential for neuroimaging findings to aid in predicting individual treatment outcomes (Gratton et al., 2020). Approaching this complex issue through multiple pathways that focus on both group differences and individual variation may be required for further progress in identifying predictors of treatment success that can be applied in clinical settings.

One potential exciting direction highlighted by our study is exploring the clinical utility of the substantial individual differences found here. While most variation was not related to group membership based on MD diagnosis or treatment response, individual variation could still be relevant in the context of MD and its treatment. Particularly the high individual variation in regions involved in cognitive control may be important, as MD has been proposed to involve abnormal functional connectivity within and among the CCN and other resting state networks (DMN and SN; Brakowski et al., 2017; Mulders et al., 2015). As both MD symptom profiles and cognitive control processes are complex and heterogenous (Fried & Nesse, 2015; Joormann & Tanovic, 2015), a more detailed exploration of such specific features in relation to individual differences in the brain may provide more insight. This approach may be useful more broadly as well, as a concern raised in neuroimaging research on mental health conditions proposes that diagnostic categories may be too broad to capture specific neurobiological processes (Blackburn, 2019; Cuthbert, 2014; Holtzheimer & Mayberg, 2011; Insel et al., 2010). A related issue is that features of the brain have been found to differ depending on the stage of the mental health condition (e.g., first episode or recurrent depression; Frodl et al., 2003; Kronmüller et al., 2009; McGorry et al., 2006; Sheline et al., 2003; Yüksel et al., 2018). Several papers have therefore called for studies examining the brain in relation to more specific symptoms and/or stages of MD (Cuthbert, 2014; Holtzheimer & Mayberg, 2011; McGorry et al., 2006). Investigating the clinical relevance of individual variation in the brain using such a fine-grained approach may help disentangle if and how individual differences relate to depression symptoms and treatment effects.

Contrary to our hypotheses, the similarity of FC across participants within time points (i.e., the effect of consistent change over time across participants) was smaller than common FC in all main analyses. We believe that this happened because, in the design of the similarity matrix for this study, time-specific FC did not include any similarity values calculated within an individual (see **Figure 1e**). Since similarity within individuals over time points was high, the fact that such similarities were not part of the average for the time-specific FC may have led to lower averages compared to the common FC similarity, which did include similarity within individuals. This is supported by the observation that the similarity estimates for time-specific FC were no longer significantly smaller in two of the exploratory analyses, where within individual similarity values were part of the average (see **Figure S5b**).

Even so, session-specific FC across participants did not contribute to the overall variance observed in the data beyond common FC, indicating changes over session did not occur the same way across participants, not even in patients receiving the same treatment. This finding is discrepant with previous research summarized in a systematic review that did identify similar changes across treatment (Gudayol-Ferré et al., 2015). Next to other methodological differences (e.g. whole-brain FC vs specific edges), this may be accounted for by the fact that within-subject designs studying changes over time typically calculate change within an individual and then examine consistencies in these changes across participants. Here instead, our main analyses examined similarities over time points across participants directly. Previous study designs more closely resemble our exploratory analyses, which did highlight a contribution of individual- and session-specific FC in patients exclusively, indicating an effect of antidepressant treatment over time. The localization of this effect (temporal poles, subgenual cingulate cortex and medial temporal areas) also partly overlapped with previous findings (Gudayol-Ferré et al., 2015; Mulders et al., 2015). Notably, our approach did not require changes to be in the same direction for each individual, instead highlighting regions where change took place within individuals regardless of direction. Exploring these individual patterns of change may help unpack the variable findings in the literature (Dichter et al., 2015; Fonseka et al., 2018).

Our study had several limitations. First, the approach we utilized only considered the relative contribution of the included sources of variance, and therefore could not indicate if additional sources may have been missed. For simplicity, we examined only a limited number of categorical variables, but future research could include additional (continuous) factors (e.g., age; Geerligs et al., 2015). Second, our study would have benefitted from longer fMRI recordings. While the amount of data we had provided acceptable reliability for whole-brain FC estimates (Gordon et al., 2017), recent studies suggest larger amounts of data (e.g., up to 200 minutes per participant) are needed to achieve high reliability of individual connectivity estimates, especially for single connections and non-cortical regions (Gratton et al., 2020). In addition, like most research in psychology, our sample and we as researchers came from ‘WEIRD’ (Western, Educated, Industrialized, Rich and Democratic) populations that represent only ∼12% of humanity (Henrich et al., 2010), indicating the need to invest in research done with and by more diverse people. Lastly, the use of biological sex and how it was measured was suboptimal. Namely, sex is not binary and other relevant, interrelated factors, such as gender expression and gender identity were not accounted for, which may have led to measurement errors (Lindqvist et al., 2021). However, given the indications of the importance of sex and gender in the context of antidepressant treatment, we decided to use the information available to us.

Here we show that common and individual-specific patterns of functional connectivity explain most of the variance in FC data, while more commonly reported group-specific patterns account for only a small amount. To our knowledge, no other studies have quantified individual variation in neuroimaging data in the context of MD and antidepressant treatment. Future studies should examine individual FC patterns and explore whether and how they relate to individual-specific symptom progression and treatment outcomes.

## Data Availability

The data used in this manuscript has been collected as part of the CAN-BIND initiative, an Integrated Discovery Program of the Ontario Brain Institute (OBI). OBI has released data from CAN-BIND's foundational study which aims to identify biomarkers that predict treatment response in people with depression. The dataset currently available on Brain-CODE includes baseline and longitudinal data from participant follow-up visits in weeks 2 to 8 of the study (Phase 1). All data have been standardized, cleaned and curated to maximize utility for analysis across different data modalities, and imaging data was converted to a BIDS-friendly naming convention. For access requirements, please visit OBI's Brain-CODE Neuroinformatics Platform (https://www.braincode.ca/) or.

## Notes

### Competing Interest Statement

Benicio N. Frey has received grant/research support from Alternative Funding Plan Innovations Award, Brain and Behavior Research Foundation, Canadian Institutes of Health Research, Hamilton Health Sciences Foundation, J. P. Bickell Foundation, Ontario Brain Institute, Ontario Mental Health Foundation, Society for Women's Health Research, Teresa Cascioli Charitable Foundation, Eli Lilly and Pfizer, and has received consultant and/or speaker fees from AstraZeneca, Bristol-Myers Squibb, Canadian Psychiatric Association, CANMAT, Daiichi Sankyo, Lundbeck, Pfizer, Servier and Sunovion. Roumen V. Milev has received consulting and speaking honoraria from AbbVie, Allergan, Janssen, KYE, Lundbeck, Otsuka, and Sunovion, and research grants from CAN-BIND, CIHR, Janssen, Lallemand, Lundbeck, Nubiyota, OBI and OMHF. Sagar Parikh has been a consultant to Takeda, Bristol Myers Squibb, Lund- beck; has had a research contract with Assurex; has equity in Mensante. Raymond W. Lam has received speaker and consultant honoraria or research funds from AstraZeneca, Brain Canada, Bristol-Myers Squibb, the Canadian Institutes of Health Research (CIHR), the Canadian Network for Mood and Anxiety Treatments, the Canadian Psychiatric Association, Eli Lilly, Janssen, Lundbeck, Lundbeck Institute, Medscape, Otsuka, Pfizer, Servier, St. Jude Medical, Takeda, the University Health Network Foundation, Vancouver Coastal Health Research Institute, Allergan, Asia-Pacific Economic Cooperation, BC Leading Edge Foundation, Healthy Minds Canada, Michael Smith Foundation for Health Research, MITACS, Myriad Neuroscience, Ontario Brain Institute, Otsuka, Unity Health, Viatris, and VGH-UBCH Foundation. Sidney H. Kennedy has received honoraria or research funds from Abbott, Alkermes, Allergan, Boehringer Ingelheim, Brain Canada, CIHR, Janssen, Lundbeck, Lundbeck Institute, Ontario Brain Institute, Ontario Research Fund, Otsuka, Pfizer, Servier, Sunovion, Sun Pharmaceuticals, and holds stock in Field Trip Health. Daniel J. Müller has received consulting and speaking honoraria from Lundbeck and Genomind. Dr Soares has received consulting and speaking honoraria from Pfizer, Otsuka, Bayer, Eisai and research grants from CAN-BIND, CIHR, OBI, and SEAMO. Stephen C Strother is a senior Scientific Advisor and shareholder in ADMdx, Inc., which receives NIH funding, and during the period of this research, he had research grants from Brain Canada, Canada Foundation for Innovation (CFI), Canadian Institutes of Health Research (CIHR), and the Ontario Brain Institute in Canada. Other authors declare no competing financial interests.

### Clinical Trial

NCT01655706

### Clinical Protocols

https://osf.io/79gv8/?view_only=9e105962ce4c4ebf8cf35393471a7b69

### Funding Statement

CAN-BIND is an Integrated Discovery Program carried out in partnership with, and financial support from, the Ontario Brain Institute, an independent non-profit corporation, funded partially by the Ontario government. The opinions, results and conclusions are those of the authors and no endorsement by the Ontario Brain Institute is intended or should be inferred. Additional funding is provided by the Canadian Institutes of Health Research (CIHR), Lundbeck, and Servier. Funding and/or in-kind support is also provided by the investigators' universities and academic institutions. All study medications are independently purchased at wholesale market values. We acknowledge support to Gwen van der Wijk in the form of a Mamdani Family Foundation Graduate Scholarship and Alberta Graduate Excellence Scholarship (AGES: International). The funders had no role in the design, analysis, interpretation, or publication of this study

### Author Declarations

Ethics approval was obtained from all participating centres. The ethics committees include: UBC Clinical Research Ethics Board (Vancouver); UCA Conjoint Health Research Ethics Board (Calgary); University Health Network Research Ethics Board (Toronto); CAM Research Ethics Board (Toronto); Hamilton Integrated Research Ethics Board (Hamilton); and QNS Health Sciences and Affiliated Teaching Hospitals Research Ethics Board (Kingston). The participants provided written, informed consent for all study procedures.

