## Supplementary materials for "Large Individual Differences in Functional Connectivity in the Context of Major Depression and Antidepressant Pharmacotherapy"

*Matching for subgroup analyses*

We matched participants to the smallest subgroup in each analysis, creating a subsample of 72 and 48 participants for the patients and controls, and patients only analyses, respectively. Two (female vs male) by 2 (controls vs patients or responders vs non-responders) ANOVAs revealed no statistically significant differences between groups in age and years of education (all *F*s < 1.67, all *p*s > .20). The number of female and male participants was equal in the matched groups, and the distributions of scanner types varied little across groups (max. difference was 4 participants).

*Results of supplementary analyses*

The analyses excluding ROIs with fewer than 8 voxels yielded highly similar results as reported in the main analyses. Namely, the main contributors to the overall variance in all three samples were the common and individual effects, while sex, MDD diagnosis, response to treatment and their interactions contributed a small but significant amount (see **Figure S3**). The matched subsamples analyses revealed the same pattern of results (**Figure S4**). This indicates that the findings in the main article were not meaningfully influenced by differences in scanner types, years of education or unequal group sizes.

**Figure S3.1**. Normalized relative effect magnitude in each region of the brain for each effect (common, response, Sex, response*sex and individual) in patients only. The response, sex and response*sex effects are presented on a narrower colour scale in **Figure S3.2** so the localization of these effects can be distinguished.

**Figure S2**. Normalized relative effect magnitude in each region of the brain for the smaller effects in patients and controls (A) and patients only (B) on a narrower colour scale.

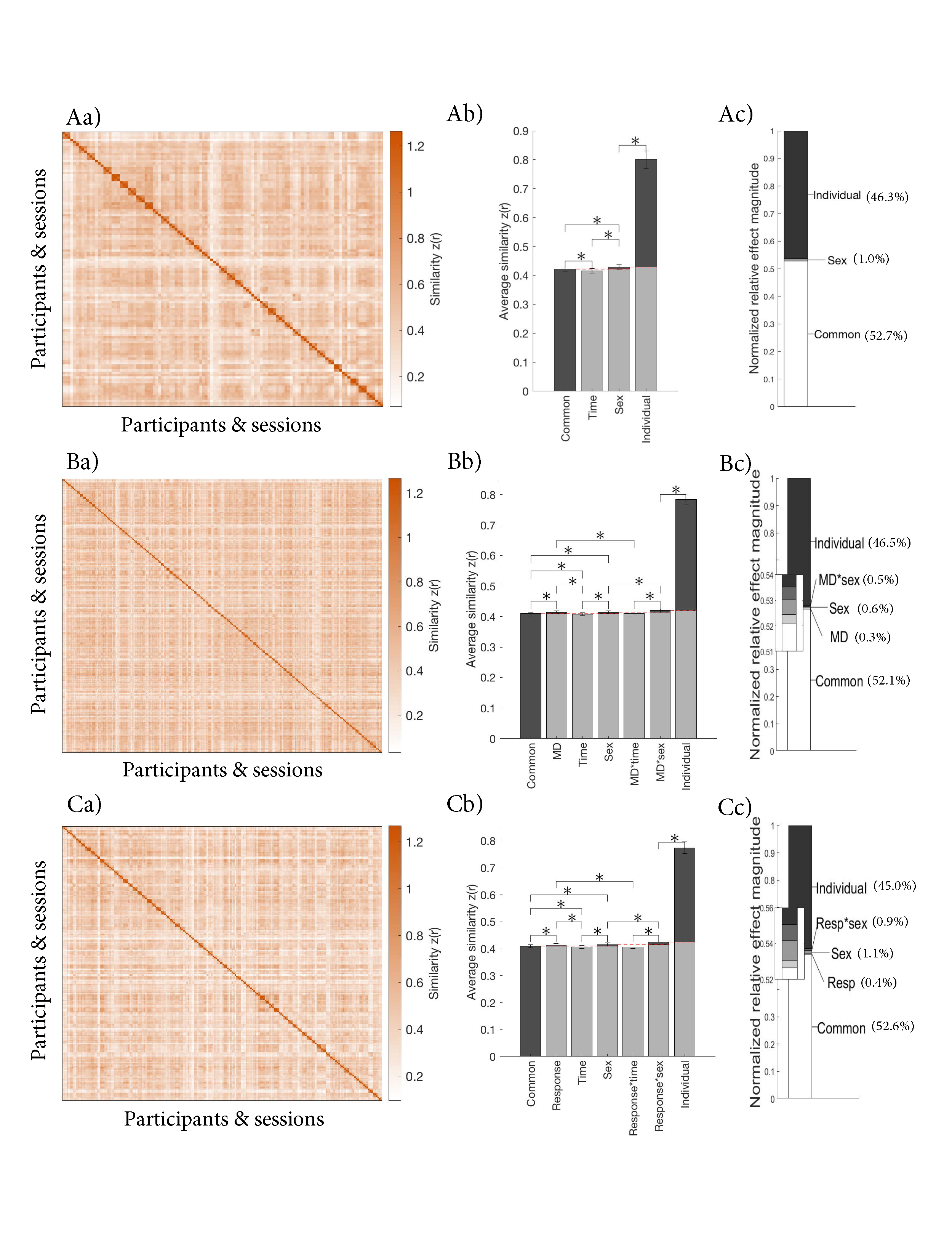

**Figure S3**. Contributions of different sources of variance in FC between ROIs with more than 8 voxels in A) controls only, B) patients and controls, and C) patients only. a) Similarity matrix displaying the Fisher transformed correlations between the whole-brain FC of different participants and sessions. The matrices are organized by group (sex and MD diagnosis or response), individual and session (baseline, week 2 and week 8) as illustrated in **Figure 1**; b) Average similarity for each effect. **Figure 1e** illustrates how each average was calculated. The red dashed lines indicate the baseline for each effect, which was subtracted to calculate the normalized relative effect magnitudes; c) Normalized relative effect magnitude for each effect.

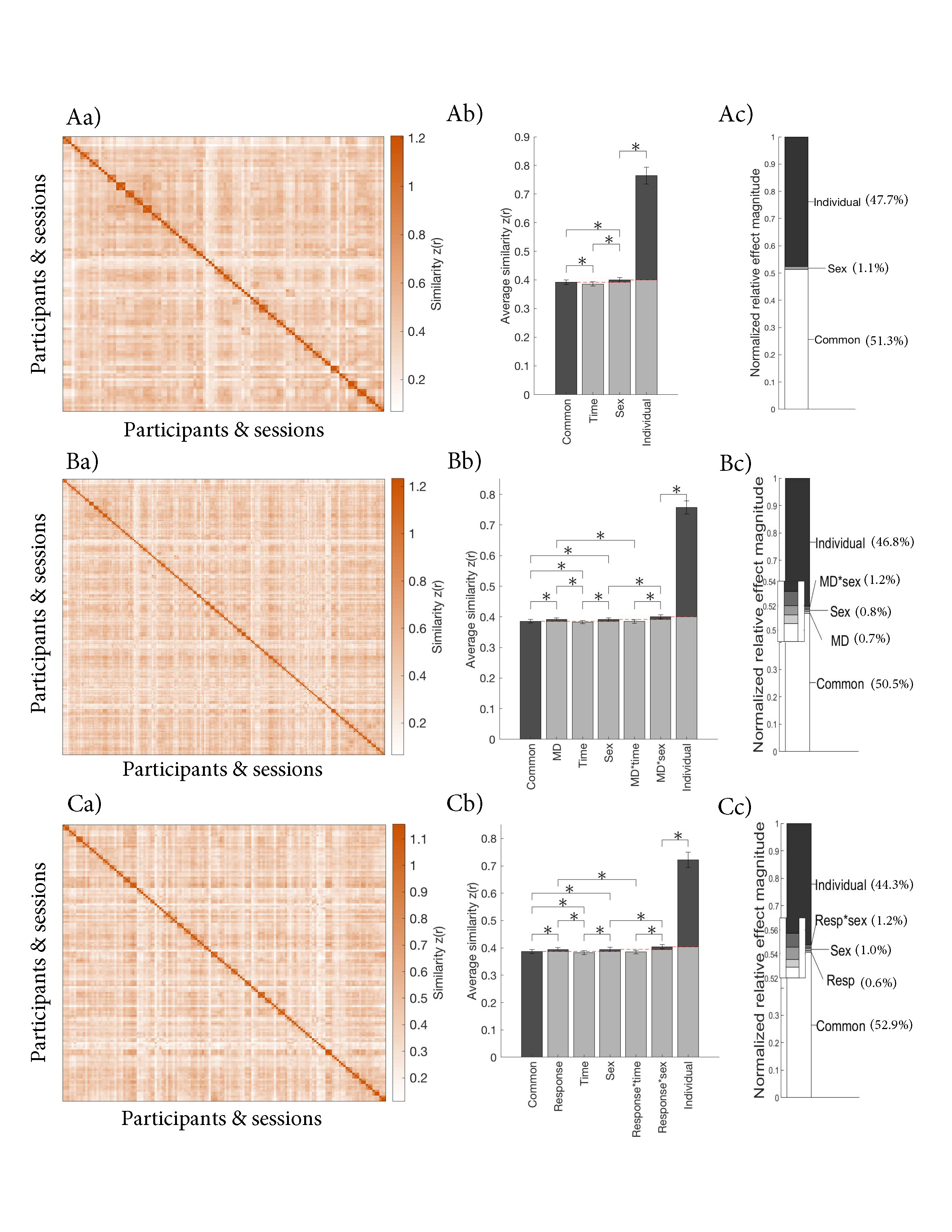

**Figure S4.** Contributions of different sources of variance in FC for samples match on scanner type, sex, age and education in A) controls only, B) patients and controls, and C) patients only. a) Similarity matrix displaying the Fisher transformed correlations between the whole-brain FC of different participants and sessions. The matrices are organized by group (sex and MD diagnosis or response), individual and session (baseline, week 2 and week 8) as illustrated in **Figure 1**; b) Average similarity for each effect. **Figure 1e** illustrates how each average was calculated. The red dashed lines indicate the baseline for each effect, which was subtracted to calculate the normalized relative effect magnitudes; c) Normalized relative effect magnitude for each effect.

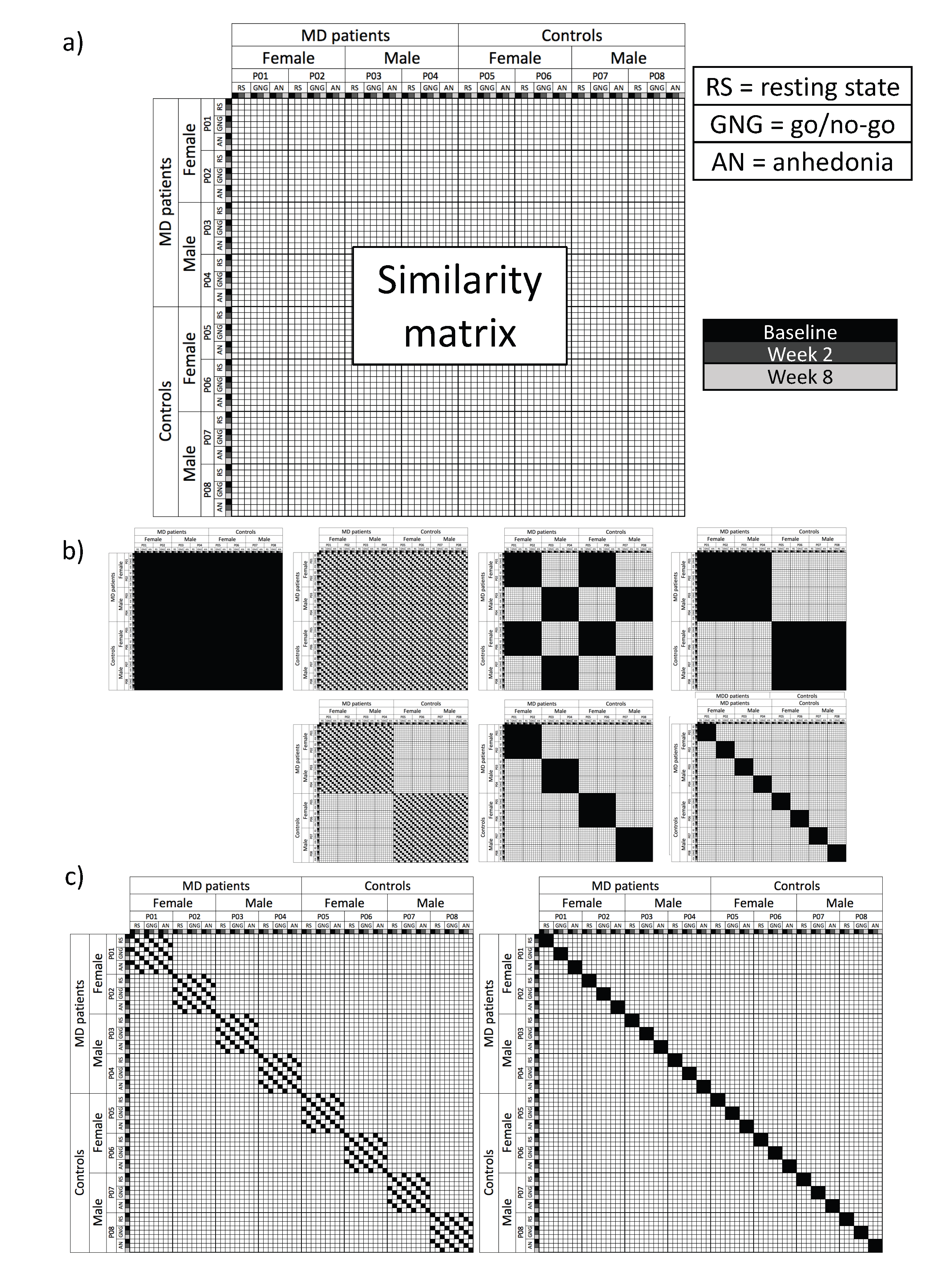

**Figure S5**. Illustrations of the similarity matrix and matrix configurations used to calculate the averages for each effect in the exploratory analyses, taking the patients and controls sample as the example. a) Structure of the similarity matrix exemplified for eight participants. There is now a separate row for each participant, session and task (instead of just for each participant and session in the main analyses). b) Matrix configuration for each of the effects that were also examined in the main analyses (see **Figure 3.1e**). The effects were calculated by averaging over the black squares in the matrix. From left to right and top to bottom, the patterns represent the (similar FC across all participants and sessions), the session effect (similar FC across participants within sessions), the sex effect (similar FC among female and among male participants), the MD effect (similar FC among patients with an MD diagnosis and among controls), the MD*session interaction (similar FC among patients and controls within sessions), the MD*sex interaction (similar FC among female patients, male patients, female controls and male controls), and the individual effect (similar FC within individuals across sessions). c) Matrix configurations of the effects only examined in the exploratory analyses. The individual*time interaction (similar FC within individual and session) is illustrated on the left, while the individual*task interaction (similar FC within individual and task) is illustrated on the right.

**Figure S6**. Normalized relative effect magnitude in each region of the brain for selected effects from the exploratory analyses (common, individual, individual*task, and individual*session) in patients and controls only.
